# The global impact of the first Coronavirus Disease 2019 (COVID-19) pandemic wave on vascular services

**DOI:** 10.1101/2020.07.16.20153593

**Authors:** The VERN COVER study collaborative, Ruth A Benson, Sandip Nandhra

## Abstract

**Background:** The Coronavirus Disease 2019 (COVID-19) pandemic is having an unprecedented impact on healthcare delivery. This international qualitative study captured the global impact on vascular patient care during the first pandemic ‘wave’.

**Methods:** An online structured survey was used to collect regular unit-level data regarding the modification to a wide range of vascular services and treatment pathways on a global scale.

**Results:** The survey commenced on 23^rd^ March 2020 worldwide. Over six weeks, 249 vascular units took part in 53 countries (465 individual responses). Overall, 65% of units stopped carotid surgery for anyone except patients with crescendo symptoms or offered surgery on a case-by-case basis, 25% only intervened for symptomatic aortic aneurysms cancelling all ‘elective’ repairs. For patients with symptomatic peripheral arterial disease 60% of units moved to an endovascular-first strategy. For patients who had previously undergone endovascular aortic aneurysm repair, 31.8% of units stopped all postoperative surveillance. Of those units regularly engaging in multidisciplinary team meetings, 59.5% of units stopped regular meetings and 39.1% had not replaced them. Further, 20% of units did not have formal personal protective equipment (PPE) guidelines in place and 25% reported insufficient PPE availability.

**Conclusions:** The COVID-19 pandemic has had a major impact on vascular services worldwide. There will be a significant vascular disease burden awaiting screening and intervention after the pandemic.

## Introduction

The Coronavirus Disease 2019 (COVID-19) has had a profound effect on the availability of surgical resources^1^.

Vascular services have been severely affected by these challenges. Some vascular societies have issued guidance on what operative case-mix should be undertaken during the pandemic^2-4^. These include adapting service provision for elective and urgent vascular presentations such as stroke and aortic aneurysm. However, the exact impact of the pandemic is still unknown^5^.

The Vascular and Endovascular Research Network (VERN) is an established vascular research collaborative^6-9^, which responded rapidly to the pandemic by delivering the **<u>CO</u>**VID-19 **<u>V</u>**ascular S**<u>ER</u>**vice (COVER) study, an international prospective mixed-methodology project. The aim of the first part of the COVER study described here (Tier 1), is to document fluctuations in vascular services globally during the first phase of the pandemic.

## Methods

International guidelines on designing and reporting of surveys were used^10^. The study protocol is available online (https://medrxiv.org/cgi/content/short/2020.05.27.20114322v1; ISRCTN: 80453162).

A remote digital survey was developed by a global team of vascular healthcare-professionals. Questions related to all aspects of vascular care, including staff availability, multidisciplinary team input, and personal protective equipment (PPE) (Appendix B). Results reported here are for the period 23rd March 2020 - 3rd May 2020, divided into three two-week periods for comparison. Duplicate responses were removed.

International/continental comparisons were performed, where possible, to describe relative change in practice. A score of 0 to 3 was allocated to each answer based on perceived relative service reduction by 12 VERN healthcare-professionals (‘0’ representing no change,’3’ representing most significant change; Appendix C).

## Results

Overall, 465 completed survey responses were collected from 249 different units in 53 countries across six continents (Appendix D). Figure 1 shows all unit responses together with overall service reductions and worldwide response. Individual countries’ reduction in service measures are available in Appendix C.

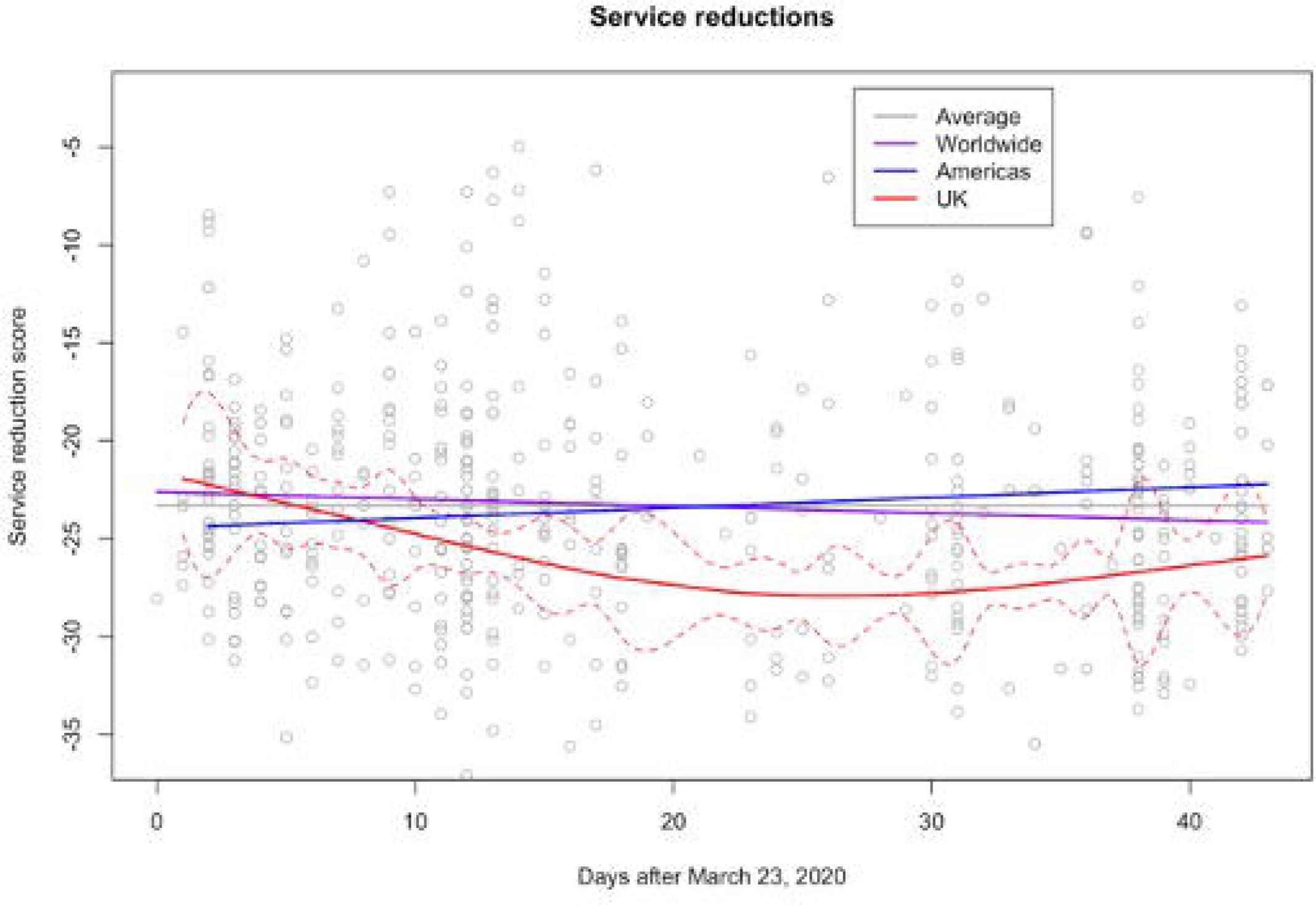

### Carotid surgery

Globally, 17.7% of units only offered intervention to patients with crescendo transient ischaemic attacks, 43.5% continued to offer surgery on a ‘case by case’ basis and 36.4% made no changes to their carotid practice.

### Aortic screening programmes

Of those units offering aortic aneurysm screening services, 45.8% stopped all screening activities; 18% continued a reduced programme and 10% continued screening as usual.

### Aortic pathologies

Thresholds for abdominal aortic aneurysm (AAA) repair were raised in the majority of centres; 11.7% of vascular units limited surgery to >6.5cm in maximal diameter, 16.4% to those >7cm, 25.1% to symptomatic or ruptured AAA and 2.3% to AAA suitable for endovascular AAA repair (EVAR) only. Despite this, 25.1% reported no change in practice. Access to EVAR out of hours was initially available to 10% of responding units, increasing to 20% in the following four weeks. Overall, only 14.2% of units maintained a 24/7 EVAR service, 26.3% maintained an ‘in hours’ service, 31.5% offered EVAR for urgent cases only and 18.5% were able to run their service on an ad hoc basis only. Post-EVAR surveillance continued as normal in 24.6% of units. However, 35.2% had reduced availability and 31.8% stopped it completely. The majority of units (56.6%) maintained their pathways for acute aortic syndromes (type B aortic dissection, penetrating aortic ulcer and intramural haematoma). A small proportion (5.9%) moved to conservative management only, whilst 4.5% were offering early endovascular surgery and 26.6% limited surgery to ruptures.

### Lower limb

Changes to the management of lower limb pathologies are shown in table 1 for each two-week period. The majority of units began offering a greater proportion of their patients major amputation or palliation rather than attempting revascularisation for chronic limb-threatening ischaemia, with many moving to an endovascular-first treatment strategy.

### Outpatient clinics

Whilst 27.5% of units moved to a triage clinic system, 29.0% cancelled all planned outpatient clinics. Use of technology permitted 14.9% of units to move to video or telephone clinics, with 18.7% including subsequent triage for attendance if required (18.7%). The use of ‘hot’ clinics (reserved for acute/urgent patients) increased during the pandemic; 79.1% of units reported using some form of hot clinic to accommodate vascular patients.

### Multidisciplinary team (MDT) meetings

Overall, 32.2% of units who normally participated in an MDT continued with face to face meetings; 59.5% stopped regular face to face meetings, and of those, 39.1% did not replace them. Overall, 36.8% moved to remote conferencing.

### Staff redeployment

Globally, 5.5% of senior surgeons were redeployed to support other specialties, compared to 53.5% of junior vascular surgical staff redeployed from the vascular team to other specialties.

### Personal Protective Equipment

The majority (80.5%) of units had PPE guidance in place; 25% did not have access to adequate PPE at the start compared to 18% at the end of this period.

## Discussion

The COVER study is the first international prospective study of unit-level vascular surgical practice during the COVID-19 pandemic. Findings from Tier 1 suggest radical changes in practice in a range of services.

One notable change across participating vascular units is the reduction in AAA screening activity. The benefit of AAA screening, and likelihood of finding a new AAA (<1.5%)^11^ must be balanced against the risk of COVID-19 transmission and allocation or resources. Given that the majority of units have reported higher size thresholds for AAA intervention, the chances of finding AAA large enough to be considered for repair at this time even lower. UK National AAA Screening Programme data suggest that 809 threshold AAAs are identified annually (2018)^12^, which implies that there will be a UK backlog of approximately 130 AAAs relating to this 6-week study period, with resource implications post-pandemic. This will be replicated to some degree worldwide.

Another very common finding is the reported preference for endovascular strategies to address aortic and peripheral arterial disease; this is thought to be based on a drive to minimise hospital stay and reduce demand on intensive care beds^13,14^. For EVAR, a paradigm has been created where potentially more EVAR is performed during the pandemic, but with a reduction in post-EVAR surveillance. There are important implications relating to the financial resources, operating time and staffing that will be required to catch up with missed scans and scheduled operations as services begin to resume. Vascular patients will be competing with the estimated 28 million operations cancelled or postponed during the pandemic peak^15^. For lower limb pathology, the results of an increased endovascular approach on limb-related outcomes will also be important to follow.

MDT meetings support individual clinician decision-making by navigating complex decisions through a multifaceted approach. COVID Guidelines have provided recommendations that potentially go against our usual surgical inclinations^2-4^. Anecdotally, patients who may have received active treatment pre-pandemic were being palliated due to the perceived high risk of intervention, especially if they tested positive for COVID-19. Strategies have moved towards endovascular management where open surgery would have been the surgeon’s usual preference. Replacing a face-to-face MDT with virtual meetings has facilitated ongoing access to MDT support for such complex decision making during this challenging period.

### Study limitations

Despite the large number of units taking part, correlating individual country or regional data with dates of lockdown is challenging. Dates of lockdown were, however, similar for countries providing the majority of responses (UK, Germany, USA). All participating units entered lockdown in March 2020, and were in lockdown when the survey began.

As the COVER study continues, we anticipate the widening of practice in the immediate future. If there are any subsequent COVID-19 ‘waves’ in areas that are ‘past the peak’, or in locations where the pandemic peak has yet to occur, this data will support vascular surgeons when deciding on how to adapt practice once again. The impact of cancellations has been modelled for general surgical procedures and predicts a large number of excess deaths^16^. It will be important for vascular surgery to have similar data available in the same way.

## ISRCTN

80453162

## Data Availability

On request.

## Acknowledgements

The VERN executive committee would like to formally acknowledge the support and collaboration with the following international groups and networks: Vascular Society of Great Britain and Ireland (VSGBI), Rouleaux Club (RC), GLOBALSurg, British Society of Endovascular Therapy (BSET), Singapore Vascular Surgical Collaborative (SingVasc), Vascupedia (European vascular education platform), VASCUNET (collaboration of Vascular Registries), Australian and New Zealand Vascular Trials Network (ANZVTN), Audible Bleeding (evidence based podcast, USA), British Society of Interventional Radiology (BSIR), BSIR Trainees (BSIRT), International Working Group in the Diabetic Foot (IWGDF), the European Society of Vascular Surgery (ESVS), European Vascular Surgeons in Training (EVST) and STARSurg (UK based student surgical research network). We would also like to thank Sonia Kandola and the Department of Research and Development at University Hospitals Coventry and Warwickshire NHS Trust, Coventry, UK, Irene Cruikshanks and Thomas Koller for Spanish and German translations. These partner groups and colleagues have enabled dissemination of the study and participation from multiple nations worldwide which has been invaluable in the success of the COVER study to date.

## Appendices

### Appendix A

#### Writing committee

Ruth A Benson, Sandip Nandhra, Joseph Shalhoub, Nikesh Dattani, Graeme K Ambler, David C Banquet.

#### Study Steering committee

Ruth A Benson and Sandip Nandhra (study co-leads). Joseph Shalhoub, Graeme K Ambler, Nikesh Dattani, David C Bosanquet, Rachael Forsythe, Sarah Onida, George Dovell, Louise Hitchman, Ryan Preece, Athanasios Saratzis (co-chief investigator), and Chris Imray (co-chief investigator).

#### International steering team

United States of America: Adam Johnson

Asia (Hong Kong SAR China/Malaysia/Singapore): Andrew Choong, Jun Jie Ng

Australia and New Zealand: Sarah Aitken, Jana-Lee Moss

#### Statistical analysis

Graeme K Ambler.

#### Tier 1 database management and quality assurance

Ruth A Benson, Sandip Nandhra, Graeme K Ambler

#### Study communications committee

Ryan Preece, Louise Hitchman, Rachael Forsythe

#### Collaborators

Abhilash Sudarsanam, Adam Tam, Adam W. Beck, Adel Barkat, Adnan Bajwa, Ahmed Elbasty, AI Awopetu, Akio Kodama, Aksim G Rivera, Alberto Munoz, Alberto Saltiel, Alejandro Russo, Alex Rolls, Alexandros Kafetzakis, Ali Kimyaghalam, Ali Kordzadeh, Amanda Shepherd, Aminder Singh, Andrea Mingoli, Andreas M. Lazaris, Andrej Isaak, Andres Marin, Andrés Reyes Valdivia, Andrew Batchelder, Andrew Duncan, Angeliki Argyriou, Anthony S Jaipersad, Antonio Freyrie, António Pereira-Neves, Anver Mahomed, Arda Isik, Arkadiusz Jawien, Asad J. Choudhry, Ashwin Sivaharan, Athanasios Giannoukas, Athanasios Papaioannou, Athanasios Saratzis, Ayman Abbas, Bakoyiannis Christos, Bekir Bogachan Akkaya, Bella Huasen, Bibombe Patrice, Mwipatayi, Bilal Azhar, Boboyor Keldiyorov, Brant W. Ullery, Carlo Pratesi, Carlos A. Hinojosa, Carlos F Bechara, Carolina Salinas Parra, Charalabopoulos Alexandros, Charlotte Bezard, Cheong Jun Lee, Chris Davies, Christian-Alexander Behrendt, Christopher Lowe, Christos D. Karkos, Chun Ling Patricia Yih, Ciarán McDonnell, Claudia Ordonez, Craig Nesbitt, Croo Alexander, Daniel Guglielmone, Daniel T Doherty, David M Riding, Davide Esposito, Denis Harkin, Dennis H Lui, Dhafer M Kamal, Diego Telve, Dimitrios Theodosiou, Domenico Angiletta, Donald Jacobs, Edward Choke, Edward D Gifford, Efthymios Beropoulis, Eftychios Lostoridis, Eleanor Atkins, Elena Giacomelli, Elpiniki Tsolaki, Emma Davies, Emma Scott, Emmanouil Katsogridakis, Ernesto Serrano, Ertekin Utku Unal, Eugenia Lopez, Eustratia Mpaili, Fabrizio Minelli, Fatemeh Malekpour, Fatma Mousa, Felicity Meyer, Felipe Tobar, Filipa Jácome, Flavia Gentile Johansson, Fred Weaver, Gabriel AB Proaño, Gabriel Sidel, Ganesh Kuhan, Gary Lemmon, George A Antoniou, George Papadopoulos, Georgios Pitoulias, Georgopoulos Sotirios, Gerardo Victoria, Gert Frahm-Jensen, Giovanni Tinelli, Giuseppe Asciutto, Gladiol Zenunaj, Gómez Vera Carlos Eduardo, Gonzalo Pullas, Grzegorz Oszkinis, Guriy Popov, Hakkı Zafer İscan, Hannah C Travers, Hashem Barakat, Hayrettin Levent Mavioglu, Hayley Moore, Ian Chetter, Ian Loftus, Ilias Dodos, Imran Asghar, Isabelle Van Herzeele, Jacopo Giordano, James Cragg, Jason Chuen, Javier Del Castillo Orrego, Jeremy Perkins, João Rocha-Neves, Jorge H. Ulloa, José Antonio Chávez, José Vidoedo, Joseph Faraj, Joseph Mills, Juan Varela, Jun Jie Ng, Jürg Schmidli, Kakavia Kiriaki, Katarzyna Powezka, Kathryn Bowser, Katy Darvall, Kenneth McCune, Ketino Pasenidou, Kevin Corless, Kevin McKevitt, Kira Nicole Long, Konstantinos G. Moulakakis, Konstantinos Roditis, Konstantinos Stavroulakis, Konstantinos Tigkiropoulos, Kristyn Mannoia, Kumar Abayasekara, Lalithapriya Jayakumar, Lasantha Wijesinghe, Laura Drudi, Lauren Shelmerdine, Leigh Ann O’Banion, Lewis Meecham, Lisa F Bennett, Lorena Grillo, Lucy Green, Lucy Wales, Luís Loureiro, Luis Mariano Palena, Luis Mariano Palena, Mahmoud MH Tolba, Manar Khashram, Manik Chana, Manuel Pabon, Marco González, Marco Virgilio Usai, Marcos Tarazona, Maria A Ruffino, Mariano Castelli, Marie Benezit, Marina Dias-Neto, Martin Malina, Martin Maresch, Martin Mazzurco, Martin Storck, Martín Veras Troncoso, Matt Popplewell, Matteo Tozzi, Matthew Metcalfe, Matti Laine, Mhammed Rawhi, Michael Ricardo, Mingzheng Aaron Goh, Mohamed Abozeid Ahmed, Mohammed Ibrahim, Mohannad Alomari, Muayyad Almudhafer, Muhammed Elhadi, Nalaka Gunawansa, Nancy Hadjievangelou, Natasha Hasemaki, Natasha Shafique, Nathan Aranson, Nicholas Bradley, Nicolas J Mouawad, Nicole C. Rich, Nikolaos Floros, Nikolaos Patelis, Nikolaos Saratzis, Nikolaos Tsilimparis, Nilson Salinas, Nishath Altaf, Oliver Friedrich, Oliver Lyons, Olivia M.B. McBride, Orestis Ioannidis, Orwa Falah, Panagiotis Theodoridis, Paolo Sapienza, Paraskevi Tsiantoula, Patrick Chong, Patrick Coughlin, Paul Bevis, Paul Carrera, Paul Dunlop, Peng Foo Wong, Pereira Albino, Peter Rossi, Petroula Nana, Philip W Stather, Pierfrancesco Lapolla, Pierre Galvagni Silveira, Prakash Saha, Pranav Somaiya, Putera Mas Pian, Rachael L Morley, Rachel Bell, Raed M Ennab, Rafael Malgor, Raffaele Pulli, Ragai Makar, Raghuram Sekhar, Rana Afifi, Raphael Coscas, Raphael Soler, Robert F Cuff, Rodney Diaz, Rodrigo Biagioni, Rosnelifaizur Bin Ramely, Rubén Rodríguez Carvajal, Sandeep Jhajj, Sara Edeiken, Sergio Benites, Sergio Zacà, Sharath Paravastu, Sharon Chan, Sharvil Sheth, Sherene Shalhub, Shiva Dindyal, Shonda Banegas, Simon Hardy, Simona Sica, Siu Chung Tam, Sivaram Premnath, Sophie Renton, Sriram Rajagopalan, Stavridis Kyriakos, Stavros Kakkos, Stefano Ancetti, Stephane Elkouri, Stephanie Lin, Stephen Wing Keung Cheng, Stylianos G. Koutsias, Tabitha Grainger, Tamer Fekry, Tamer Ghatwary Tantawy, Tamim Siddiqui, Taohid Oshodi, Tasleem Akhtar, Thomas James Hardy, Thomas Kotsis, Thushan Gooneratne, Timothy Rowlands, Tina U. Cohnert, Tom Wallace, Tristan R A Lane, Umberto Marcello Bracale, Usman Cheema, Uzma Sadia, Vanessa Rubio, Victor Canata, Vincent Jongkind, Vipul Khetarpaul, Virginia Summerour, Walter Dorigo, Wissam Al-jundi, Xun Luo, Yamume Tshomba, Yvis Gadelha Serra.

### Appendix B

#### Tier 1 survey questions

> “Please tell us your vascular unit/ hospital/institution and city”
>
> “In what country do you work?’“
>
> “Unit / Hospital / Institution”
>
> “State / County (if applicable)”
>
> “City”
>
> “Have you filled this survey in before?”
>
> “Have you modified the working pattern for consultants/attending/faculty “
>
> “within your unit? (clinics / MDT; etc will be covered later in the survey)”
>
> “Have you or members of your junior team been asked to cross cover other surgical specialties”
>
> “How many vascular Consultants are there in your centre?”
>
> “How many vascular specialist registrars/trainees or equivalent middle grades does your centre have? If outside of the UK - ST refers to year of training e.g. ST3 is someone who is in the 3rd year of training in surgery. PGY: Post graduate year”
>
> “Have you modified the outpatient clinics within your unit?”
>
> “Are you running an emergency or hot clinic for urgent referrals?”
>
> “Are you participating in a face-to-face MDT?”
>
> If you are not running a face-to-face MDT how have you replaced this?”
>
> “Is a vascular scientist/duplex ultrasound service currently available at your centre?”
>
> “What is your centre’s usual primary cross-sectional imaging service?”
>
> “Is your primary cross-sectional imaging service available as normal?”
>
> “If you have an AAA screening programme, is this still running as normal?”
>
> “Do you still have a service running to image patients after an “
>
> “Endovascular aortic repair (EVAR) repair ? (e.g.: annual CT)”
>
> “Is there a full endovascular aortic service available?”
>
> “Are you relying on an increased endovascular strategy service first for Chronic Limb threatening Ischaemia (CLTI)?”
>
> “Do you have vascular specific inpatient beds?”
>
> “Approximately how many vascular specific inpatient beds does your unit “ “normally have?
>
> “How many vascular specific inpatient beds does your unit currently have?”
>
> Have you changed your operative practice for elective AAA surgery?”
>
> “In general have you changed your CLTI revascularisation strategy?”
>
> “In general, have you altered how you manage symptomatic carotid disease?
>
> “In general, have you modified your acute aortic syndrome (type B aortic dissection etc.)”
>
> “Do you have access to a dedicated vascular surgery list daily?”
>
> “If yes - is this running at normal capacity? Any changes to staffing (e.g. theatre team, anaesthetic cover?)”
>
> “If you had access to a hybrid theatre before the pandemic, do you still have normal access to it now?”
>
> “Has your centre disseminated a PPE policy to members of your vascular team / unit?” “Are you able to follow the policy?”
>
> “Have you got something else to add e.g. a story from your hospital or any comment?”
>
> “If you have completed the survey before, has anything changed at your centre since you last filled it in? “

#### Study Design and methodology

An online survey (SurveyMonkey^®^) was developed by the COVER study team, with vascular surgeons at junior and senior levels from the USA, UK, Australia and Singapore, to provide clear language and questions that were applicable to global practice assessment.

The survey was piloted amongst the stakeholders in the first instance to ensure language and questions were clear and appropriate.

In the first fortnight, closed and open questions were used enabling each centre to provide free-text for feedback based on the local challenges. The questions related to centres’ provision of common vascular services, imaging, screening, staff availability, theatre suite availability, multi-disciplinary team input, clinics and PPE. After one week, a preliminary review of responses to open questions (marked as ‘other’) was used to provide more closed questions, suitable for the global participants, and to support longitudinal data comparison. Centres were asked, through regular repeated advertisement via social media, e-newsletters, and established international collaborative networks, to complete the survey regularly (at least weekly).

#### Data cleaning

The raw survey data was carefully scrutinised and cleaned prior to analysis. Duplicate responses (defined as responses from the same unit on the same day), and responses which contained no usable data (e.g. where the responder had entered no more than the name and/or size of the unit without answering any of the questions about service provision) were removed. There were a number of responses where the respondent had selected ‘other’ but then typed a free-text response which corresponded with one of the pre-specified options.

These options were allocated to specific answers so that they could be counted along with the other options. These were almost exclusively responses made in the first week after the survey went live, before the number of pre-specified options was increased.

### Appendix C

#### International/continental comparison analysis

We performed international/continental comparisons, to describe relative change in practice from normal. This was achieved by allocating a score of 0/1/2/3 to each possible answer for each service evaluation question. A score was allocated based on the perceived relative service reduction (with ‘0’ representing no change and ‘3’ representing the most significant change). For example, for the question: “have you changed your operative practice for elective AAA survey?” the answer ‘no change to practice’ automatically scored 0, whereas the answer ‘limiting surgery to >7cm asymptomatic AAA’, a significant change, could be scored 1/2/3. A score for each survey question answer was independently provided by 12 COVER team members (all vascular specialists). The mean value from these responses was then used to quantify the overall change in vascular service provision for each responding unit. Centre responses were then plotted with smoothing splines used to fit the trend in the average response, and Jackknife residuals used to generate approximate 95% confidence intervals for the change in average responses over time (where there was an apparent change in responses over time). Generalised cross-validation was used to automatically choose optimal smoothing parameters.

The scores given to each answer are shown below. ‘*’ indicates responses for which a pre-specified score was mandated.

AAA = abdominal aortic aneurysm, CCU = coronary care unit, CLTI = chronic limb threatening ischemia, HDU = high dependency unit, ITU = intensive care unit, MDT = multidisciplinary team, ST.DEV = standard deviation, TEVAR = thoracic endovascular aortic repair, TIA = transient ischemic attack.

#### Reduction to services scoring data

**Table A1:** VERN Executive Committee member average scores (n=12) for COVER Tier 1 question responses when asked about the perceived significance of each response in terms of service reduction/change:

| QUESTION & HEADLINE RESPONSE | SPECIFIC RESPONSE | MEAN | ST.DEV |
| --- | --- | --- | --- |
| **Have you modified the working pattern for consultants/attending/faculty within your unit?** |  |  |  |
| YES | Consultants asked to support other specialties | 2.58 | 0.51 |
|  | Reduced elective activity | 1.75 | 0.84 |
|  | Urgent intervention only | 2.25 | 0.62 |
|  | Doubled up on call / shadow on call | 1.54 | 0.71 |
|  | Reduced number of consultants in hospital | 1.38 | 0.71 |
| NO* |  | 0.00 | 0.00 |
| **Have you or members of your junior team been asked to cross cover other surgical specialties** |  |  |  |
| YES | Yes, to medical specialties | 2.42 | 0.80 |
|  | Yes, to the emergency department | 2.25 | 0.75 |
|  | Yes, to ITU / HDU | 2.17 | 0.72 |
|  | Yes, to surgical specialties | 1.17 | 0.39 |
| NO* |  | 0.00 | 0.00 |
| **Have you modified the outpatient clinics within your unit?** |  |  |  |
| YES | Video/Telephone consultation for all | 1.83 | 0.58 |
|  | All cancelled | 3.00 | 0.00 |
|  | Video/Telephone consultation for triaged patients only | 2.21 | 0.58 |
|  | Triage of referrals and then review those eligible in person | 1.33 | 0.49 |
| NO* |  | 0.00 | 0.00 |
| **Are you running an emergency or ‘hot’ clinic for urgent referrals?** |  |  |  |
| YES | Yes, 1 - 2 days per week | 1.36 | 0.79 |
|  | Yes, 3 - 4 days per week | 1.64 | 0.67 |
|  | ≥5 per week | 2.00 | 1.26 |
| NO* |  | 0.00 | 0.00 |
| **Are you participating in a face-to-face MDT?** |  |  |  |
| YES* |  | 0.00 | 0.00 |
| NO* |  | 1.00 | 0.00 |
| *If you are not running a face-to-face MDT how have you replaced this?* |  |  |  |
|  | Not being replaced* | 3.00 | 0.00 |
|  | Video conference / teleconference | 1.55 | 0.52 |
|  | Limited Core team attending, others dialling in as required | 1.55 | 0.69 |
| **Is your primary cross-sectional imaging service available as normal?** |  |  |  |
| YES* |  | 0.00 | 0.00 |
| NO |  | 2.58 | 0.65 |
| **If you have a AAA screening programme, is** |  |  |  |
| **this still running as normal?** |  |  |  |
| YES* |  | 0.00 | 0.00 |
| NO | No, Stopped | 2.67 | 0.49 |
|  | No, Reduced | 1.58 | 0.51 |
| (Do not have a screening programme*) |  | 0.00 | 0.00 |
| **Do you still have a service running to image patients after an EVAR?** |  |  |  |
| YES* |  | 0.00 | 0.00 |
| NO | No, Stopped | 2.42 | 0.51 |
|  | No, reduced | 1.33 | 0.49 |
| **Is there a full endovascular aortic service available?** |  |  |  |
| YES | Running as normal in hours | 1.18 | 0.82 |
|  | Running as normal 24hours/day* | 0.00 | 0.00 |
|  | Yes but Ad hoc | 1.63 | 0.54 |
|  | Reduced due to COVID related sickness/absence of staff | 2.33 | 0.66 |
| NO | No (only for urgent cases) | 2.54 | 0.47 |
| **Are you relying on an increased endovascular strategy service first for CLTI?** |  |  |  |
| YES* |  | 1.00 | 0.00 |
| NO* |  | 0.00 | 0.00 |
| **Do you have vascular specific inpatient beds?** |  |  |  |
| YES* |  | 0.00 | 0.00 |
| NO* |  | 1.00 | 0.00 |
| **% fall in inpatient vascular beds (compared to normal)** |  |  |  |
|  | 0 - 25% | 0.96 | 0.85 |
|  | 26 - 50% | 1.83 | 0.61 |
|  | 51 - 75% | 2.46 | 0.50 |
|  | 76 - 100% | 2.75 | 0.62 |
| **Have you changed your operative practice for elective AAA surgery?** |  |  |  |
| YES | Symptomatic/ruptured only | 2.92 | 0.29 |
|  | Yes, >6.5cm Asymptomatic | 2.04 | 0.62 |
|  | >7cm Asymptomatic | 2.59 | 0.66 |
|  | EVAR only (i.e. only EVAR if anatomically suitable) | 2.42 | 0.64 |
| NO* |  | 0.00 | 0.00 |
| **In general, have you changed your CLTI revascularisation strategy?** |  |  |  |
| YES | Tissue loss only | 2.50 | 0.80 |
|  | Rest pain or worse | 1.92 | 0.74 |
|  | Increased endovascular strategy | 1.79 | 0.75 |
|  | More conservative decision making | 2.08 | 0.79 |
| NO* |  | 0.00 | 0.00 |
| **In general, have you altered how you manage symptomatic carotid disease?** |  |  |  |
| YES | Crescendo TIA only | 2.58 | 0.51 |
|  | Case-by-case | 1.71 | 0.81 |
| NO (still following national/international |  | 0.00 | 0.00 |
| guidelines) * |  |  |  |
| **In general, have you modified your acute aortic syndrome pathway (type B aortic dissection etc.)?** |  |  |  |
| YES | Conservative management, unless ruptured | 2.00 | 0.67 |
|  | Conservative management in a non-ITU/CCU bed | 2.42 | 0.64 |
|  | Early TEVAR and discharge | 1.96 | 0.92 |
| NO* |  | 0.00 | 0.00 |
| **Do you have access to a dedicated vascular surgery list daily?** |  |  |  |
| YES | Elective vascular lists* | 0.00 | 0.00 |
|  | Reduced but still some during the week | 1.50 | 0.50 |
|  | Urgent theatre cases only | 2.75 | 0.45 |
| NO vascular specific slots/lists |  | 2.42 | 0.50 |
| *If YES - is this running at normal capacity?*<br>*Any changes to staffing (e.g. theatre team, anaesthetic cover)?* |  |  |  |
| Running at normal capacity/no changes* |  | 0.00 | 0.00 |
| Any change specified in free text response* |  | 1.00 | 0.00 |
| **If you had access to a hybrid theatre before the pandemic, do you still have normal access to it now?** |  |  |  |
| YES | Normal access* | 0.00 | 0.00 |
|  | Reduced access | 1.83 | 0.58 |
| NO | No access at all | 2.83 | 0.39 |

**Table A2.** details the countries that provided more than 10 responses over the survey period and their mean reductions to service (total reduction Mean 23.3, max 37.2, theoretical maximum 47.2)

| **Country Name** | **Continent** | **Responses** | **Units** | **Mean** | **Std.Dev** |
| --- | --- | --- | --- | --- | --- |
| Germany | Europe | 16 | 12 | -17.432 | 5.838 |
| Colombia | Americas | 18 | 14 | -24.698 | 3.736 |
| Italy | Europe | 28 | 15 | -21.707 | 4.474 |
| Greece | Europe | 39 | 17 | -24.008 | 4.057 |
| United States | Americas | 55 | 35 | -23.526 | 4.246 |
| United Kingdom | Europe | 161 | 54 | -25.222 | 5.835 |

### Appendix D

**Table A3:**
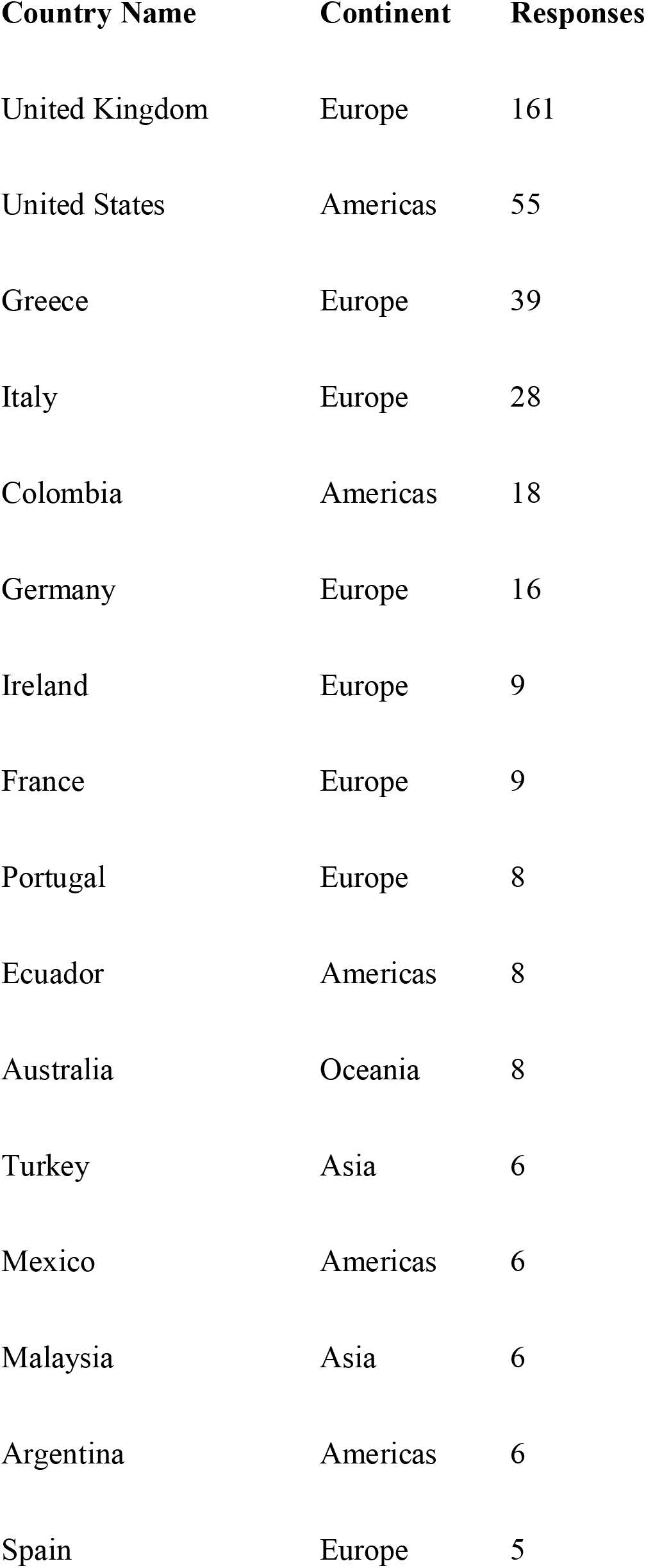
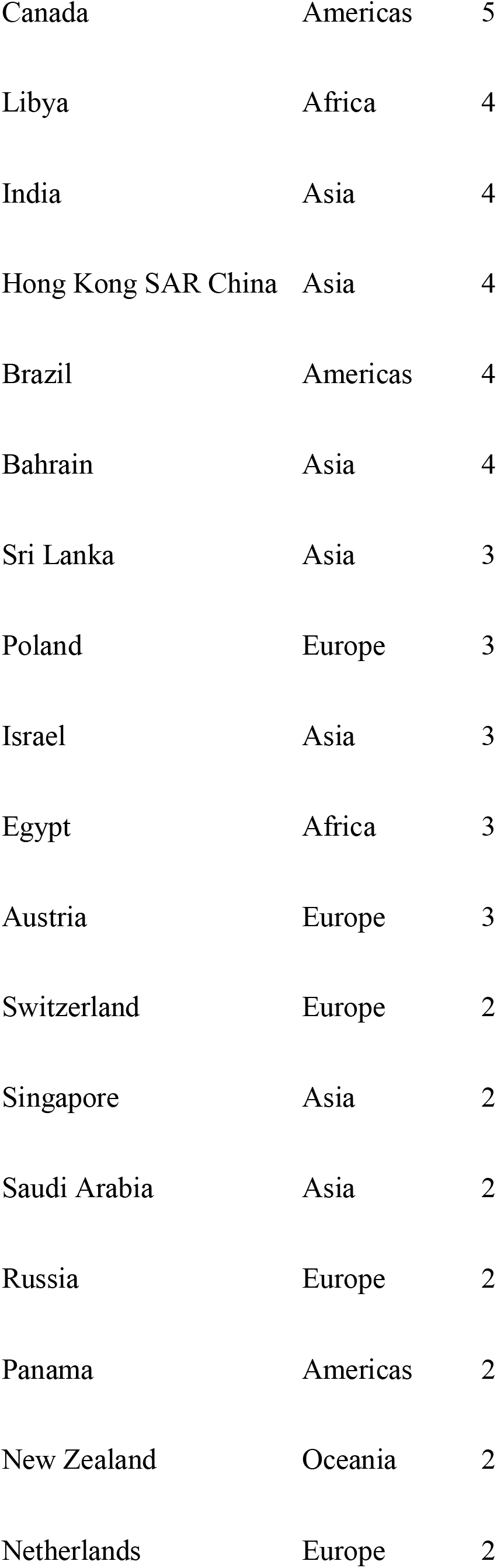
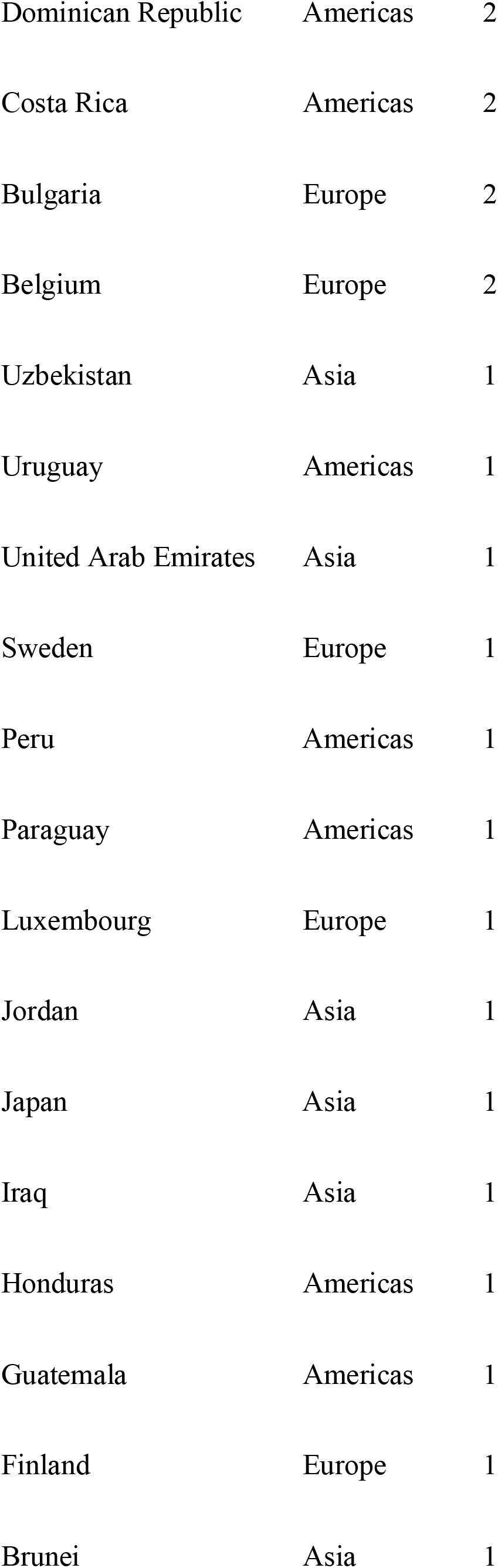
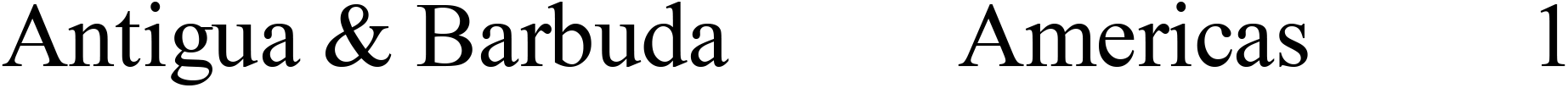
Full list of Countries surveyed and number of responses:

